# The impact of COVID-19 on Italy Web users: a quantitative analysis of regional hygiene interest and emotional response

**DOI:** 10.1101/2020.08.09.20171181

**Authors:** Alessandro Rovetta, Lucia Castaldo

## Abstract

**Background:** Between the end of February and the beginning of June 2020, Italy was certainly one of the worst affected countries in the world by the COVID-19 pandemic. During this period, Web interest in the novel coronavirus underwent a drastic surge.

**Objective:** The aim of this study was to quantitatively analyze the impact of COVID-19 on Web searches related to hygiene-preventive measures and emotional-psychological aspects as well as to estimate the effectiveness and limits of online information during an epidemic. We looked for significant correlations between COVID-19 relative search volumes and cases per region to understand the interest of the average Italian Web user during international, national, and regional COVID-19 situations. By doing so, from the analysis of Web searches, it will be possible to deduce the mental and physical health of the population.

**Methods:** To conduct this research, we used the “Google Trends” tool, which returns normalized values, called “relative search volumes” (*RSV*s), ranging from 0 to 100 according to the Web popularity of a group of queries. By comparing the *RSV*s in periods before and after the outbreak of the novel coronavirus in Italy, we derived the impact of COVID-19 on the activity of Italian netizens towards novel coronavirus itself, specifically regarding hygiene, prevention, and psychological well-being. Furthermore, we calculated Pearson’s correlations *ρ* between all these queries and COVID-19 cases for each region. We chose a *p*-value (*p*) threshold *α* = .1.

**Results:** After the two initial spikes that occurred on February 23 and March 9, 2020, the general Web interest in COVID-19 in Italy waned, as did the correlation with the official number of cases per region (*p* < .1 only until March 14, 2020). However, Web interest was similarly distributed across the regions (*ASV* = 92, *SD* = 6). We also found that all trends depend significantly on the number of COVID-19 cases at the national but not international or regional levels. Between February 20 and June 10, 2020, Web interest relating to hygiene and prevention increased by 116 and, 901*%* respectively, compared to those from January 1 to February 19, 2020 (95% *CIs:* [115.3, 116.3], [850.3, 952,2]). Significant correlations between regional cumulative Web searches and COVID-19 cases were found only between February 26 and March 7, 2020 (*ρ_best_* = .43, 95% *CI:* [.42, .44],*p* = .07). During the COVID-19 pandemic until June 10, 2020, national Web searches of the generic terms “fear” and “anxiety” grew by 8% and 21%, respectively (95% *CIs:* [8.0,8.2], [20.4,20.6]), compared to those of the period of January 1, 2018 – December 29, 2019. We found cyclically significant correlations between negative emotions related to the novel coronavirus and COVID-19 official data.

**Conclusions:** Italian netizens showed a marked interest in the COVID-19 pandemic only when this became a direct national problem. In general, Web searches have rarely been correlated with the number of cases per region; we conclude that the danger, once it arrived in the country, was perceived similarly in all regions. We can state that the period of maximum effectiveness of online information in relation to this type of situation is limited to 3-4 days from a specific key event. If such a scenario were to occur again, we suggest that all government agencies focus their Web disclosure efforts over that time. Despite this, we found cyclical correlations with Web searches related to negative feelings such as anxiety, depression, fear, and stress. Therefore, to identify mental and physical health problems among the population, it suffices to observe slight variations in the trend of related Web queries.

## Introduction

Owing to the introduction of tools for the analysis of query popularity such as Google Trends, the Internet has become one of the most important platforms for obtaining valuable data on users’ health and interests [1, 2]. Over time, an increasing number of authors have utilized Google Trends and the techniques of use have been refined by creating tutorials articles [3]. Alongside the increasing number of users who use the Web every day to obtain information on any topic, there is a growing need to control the circulation of this information to prevent fake news from affecting public health and the economy. For this reason, new branches of science called *infodemiology* and *infoveillance* were born [4]. Given the COVID-19 emergency, these two disciplines will provide useful data to researchers to evaluate the impact of the pandemic on people’s lives and welfare, the spread of fake news and infodemic monikers, as well as people’s attitudes towards an unprecedented international issue.

Between the end of February and the beginning of April, Italy was the second most affected country by COVID-19, both in terms of the total number of infected and deaths, becoming the epicenter of a shifting pandemic [5]. As evidenced in other studies, an enormous amount of information regarding the novel coronavirus has circulated in the country [6]. Although most of these were moderately infodemic, there was great interest in hygiene and prevention measures. However, no research in the scientific literature available has quantitatively analyzed the impact of COVID-19 on psychology- or hygiene-related Web searches by looking for correlations with the number of cases per region. Since, as shown in other studies, the impact of COVID-19 on the mental and physical health of the Italian population has been devastating, we believe it is of fundamental importance to study the relationship between the trend of health-related queries and the real need for assistance [7]. In fact, this could help scientists estimate the well-being of the population from the trend of Web searches.

## Methods

We used the Google Trends tool to investigate Italian netizens’ Web interest in the COVID-19 pandemic. As explained in other studies of this type, Google Trends provides normalized values, called a relative search volume (RSV), ranging from 0 to 100 in proportion to the popularity of the queries [3]. We searched for specific keywords that recorded high RSVs, in conjunction with “related queries” and “related topics”, and utilizing the results of another study on the COVID-19 infodemiology in Italy [6]. We focused on three categories of queries: generic news, hygiene, and those related to stress and anxiety. After collecting the data, we looked for correlations with the official data on COVID-19 provided by the Italian Civil Protection Department regarding the number of infected, hospitalized, dead, healed, and tested [8]. When the trends showed sufficient regularity, we looked for interpolating functions that represented them.

### Statistical analysis

We collected all the data day-by-day from February 24 to June 10, 2020. In order to have a graphical representation of regional Web search trends, we exploited this strategy by calling *RSV_i_* the Google Trends relative search volume of a specific keyword group for the region *i* and *RSV_tot_* the national relative search volume for the same keyword group, we introduced a variable *x* and imposed 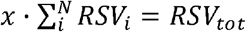, obtaining 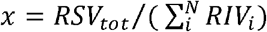. Then, we calculated a weighted relative search volume *WRSV_i_ = x·RSV_i_* for each region *i*. After that, we calculated Pearson’s correlations *ρ_ij_* between the daily *RIVs* and the number of active COVID-19 cases, total cases, new cases, home isolations, recovered, new deceased, and total deceased, for all regions. We repeated the same procedure with cumulative values in the same time interval. The regions investigated were *N* = 19. We chose *α* = .1 (10*%*) as the p-value (*ρ*) significance threshold. We also reported the thresholds *ρ_a_* for *α* = .05 (5*%*) and *α* = .01 (1*%*): *ρ_ol_* = .58, *ρ_os_* = .46, *ρ_l_* = .39. We calculated the average search volume (*ASV*) values as the average values of *RSV*s at time intervals specified in the results. For each *ASV* we reported a Gaussian 95*%* confidence interval using the formula 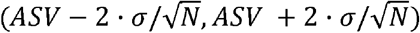 where *σ* is the standard deviation. We denote *ρ_best_* the arithmetic mean of specific groups of correlations shown in the results. To estimate the confidence interval, we used the error propagation formula 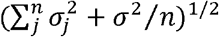, where *σ_j_* (*SEM*) is the standard deviation in the Gaussian distribution *G*(*ASV_j_, SEM_j_*). We used “Igor Pro 6.3.7” and “Microsoft Excel 2019” software for interpolations and data processing. We calculated percentage discrepancies using the formula *∆_%_* = (*y − x*)*/x·*· 100. We have calculated all errors on the functions used through the propagation formula of standard errors [9]. For all interpolations and some data, we reported the best values and the relative standard deviations using the abbreviation *SD*; in such cases, we have kept all possible significant figures provided by the software.

### COVID-19-related significant keywords

Generic news keywords: coronavirus. Category: All.

Health keywords: amuchina, disinfectant, ffp2, ffp3, gel, gloves, mask, masks, sanitizing. Category: All.

Negative emotional response keywords: anxiety coronavirus, depression coronavirus, fear coronavirus, fear covid, stress coronavirus Category: All.

### Legend

COD: COVID-19 official data, i.e. data provided by the Ministry of Health and Civil Protection.

Critical threshold: limit of daily COD beyond which there is a sudden increase in COVID-19 related interest.

Deceased: cumulative total number of deaths from COVID-19. Discharged recovered/discharged and healed: cumulative total number of patients recovered from COVID-19.

Home isolated: people currently in home-isolation due to COVID-19 infection.

Hospitalized with symptoms: people currently hospitalized for COVID-19 who exhibit symptoms.

Intensive care: people currently hospitalized for COVID-19 in intensive care.

New cases / new infected: daily increase in currently active cases of COVID-19.

New deceased: daily increase in the number of deaths from COVID-19.

Swabs: cumulative total number of COVID-19 tests performed.

Total active cases: total number of people currently infected with COVID-19.

Total cases: cumulative total number of COVID-19 cases.

Total hospitalized: cumulative total number of people hospitalized for COVID-19.

Δ total active cases / Δ total infected: daily increase in total cases of COVID-19.

## Results

### Generic news Web interest

The Web interest of Italian users towards COVID-19 was similarly distributed among all regions (*ASV* = 92, *SD* = 9) and dropped over time [Table 1, Figure 1]. Its trend, after the last peak on March 11, until June 10, 2020, is well represented by the exponential function 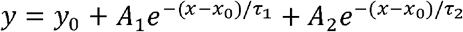 with *y*_0_ *= −*5.017, *SD* = 23.5; *A*_1_ *=* 56.801, *SD* = 10.6; *τ*_1_ *=* 0.012831, *SD* = 0.0122; *A*_2_ *=* 28.665, *SD* = 13.6; *τ*_2_ *=* 0.0995501, *SD* = 0.0473. The arithmetic mean of all the average daily COD correlations between February 24 and June 10, 2020, is not statistically significant (*ρ_best_* = .17, 95% *CI:* [.15, .19], *p* = .48). We found significant positive daily COD correlations at the end of February with hospitalized, home isolated, and recovered patients as well as with new, active, and total cases, and new and total deaths (*ρ_best_* = 45, 95% *CI:* [.44, .46], *p* = .05). After that, other substantial positive COD correlations occurred only two times [Figure 2]. We found significant negative daily COD correlations from March 8 onwards; namely with the number of COVID-19 swab tests reached several significant negative peaks between May and June, 2020 (*ρ_best_ = -*.48, 95% *CI:* [*−*.51, *−*.45], *p* = .04). The cumulative COVID-19 Web searches – COD correlations maintained significant values until around March 14, although these also showed a declining trend [Figure 3]. In fact, excluding the items “recovered”, “Δ total infected”, and “swabs”, a function representative of this correlation (February 24 – June 10, 2020) is *y = A + B ·* (1 *+ e*_(_*_C−_*_x)_*_/D_*), with *A* = 0.4360, *SD* = 0.0202; *B = −*0.34409, *SD* = 0.022; *C* = 23.438, *SD* = 1.26; *D* = 5.8573, *SD* = 1.02.

**Table 1.** COVID-19-related generic news, hygiene, prevention, and negative emotions RSVs. RSV = relative search volume, ASV = average search volume, SD = standard deviation. *South Tyrol is included.

| Region | Generic News RSV | Hygiene RSV | Prevention RSV | Negative Emotions RSV |
| --- | --- | --- | --- | --- |
| Abruzzo | 95 | 91 | 90 | 88 |
| Aosta | 99 | 68 | 83 | 0 |
| Apulia | 87 | 96 | 76 | 92 |
| Basilicata | 98 | 100 | 85 | 0 |
| Calabria | 97 | 93 | 82 | 0 |
| Campania | 83 | 93 | 75 | 94 |
| Emilia-Romagna | 96 | 81 | 85 | 100 |
| Friuli-Venezia Giulia | 95 | 81 | 85 | 0 |
| Lazio | 90 | 86 | 84 | 86 |
| Liguria | 92 | 94 | 98 | 67 |
| Lombardy | 96 | 81 | 92 | 93 |
| Marche | 91 | 85 | 86 | 79 |
| Molise | 87 | 97 | 90 | 0 |
| Piedmont | 98 | 79 | 93 | 96 |
| Sardinia | 97 | 96 | 100 | 88 |
| Sicily | 82 | 89 | 72 | 81 |
| Trentino-Alto Adige* | 80 | 68 | 59 | 0 |
| Tuscany | 95 | 83 | 90 | 70 |
| Umbria | 100 | 84 | 84 | 0 |
| Veneto | 86 | 78 | 88 | 75 |
| **ASV** | **92** | **86** | **85** | **55** |
| **SD** | **6** | **9** | **9** | **41** |

**Figure 1.**
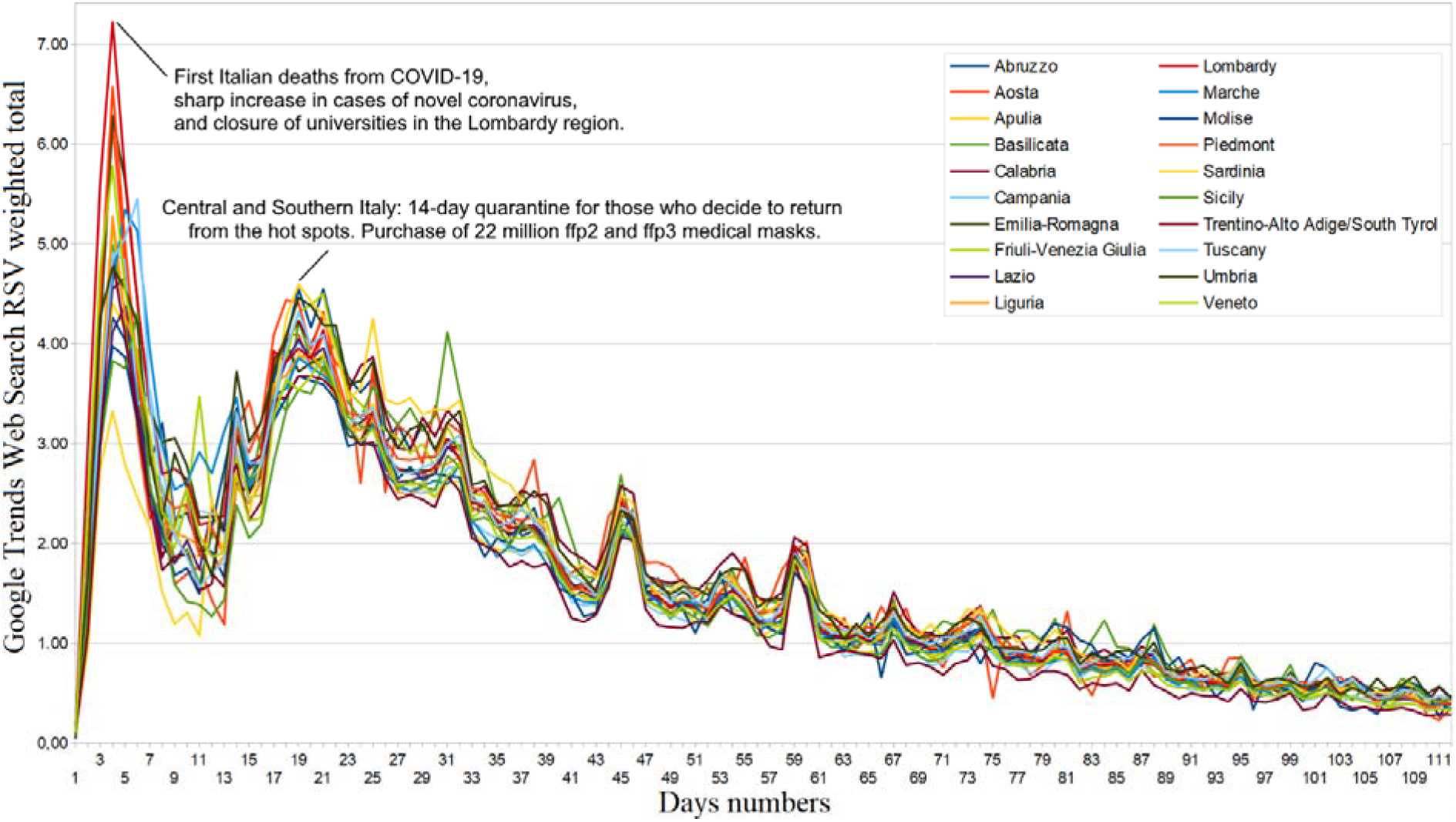
COVID-19 regional Web interest from February 20 to June 10, 2020.

**Figure 2.**
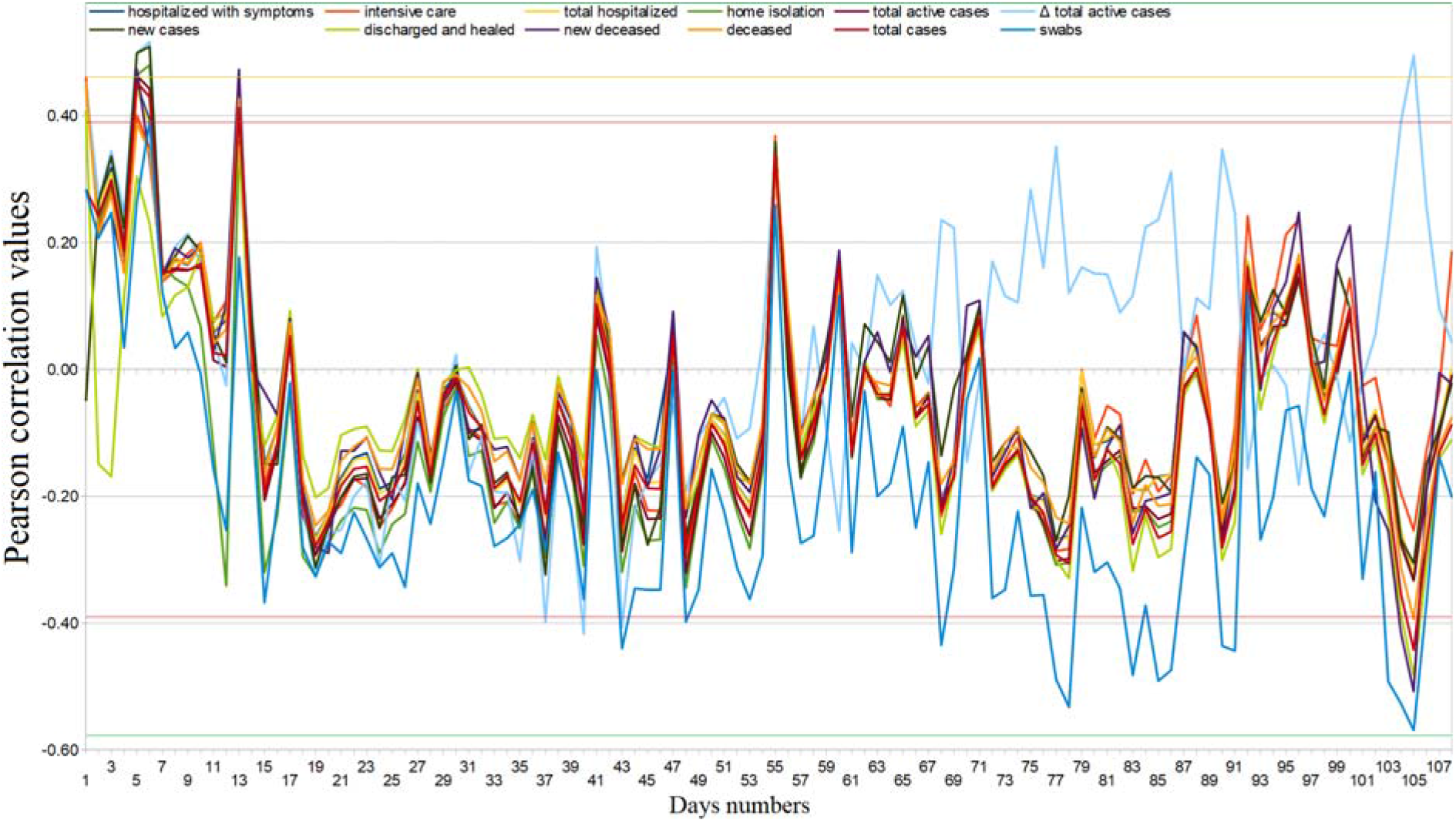
COVID-19 regional daily Web interest – official data Pearson’s correlation trends from February 24 to June 10, 2020.

**Figure 3.**
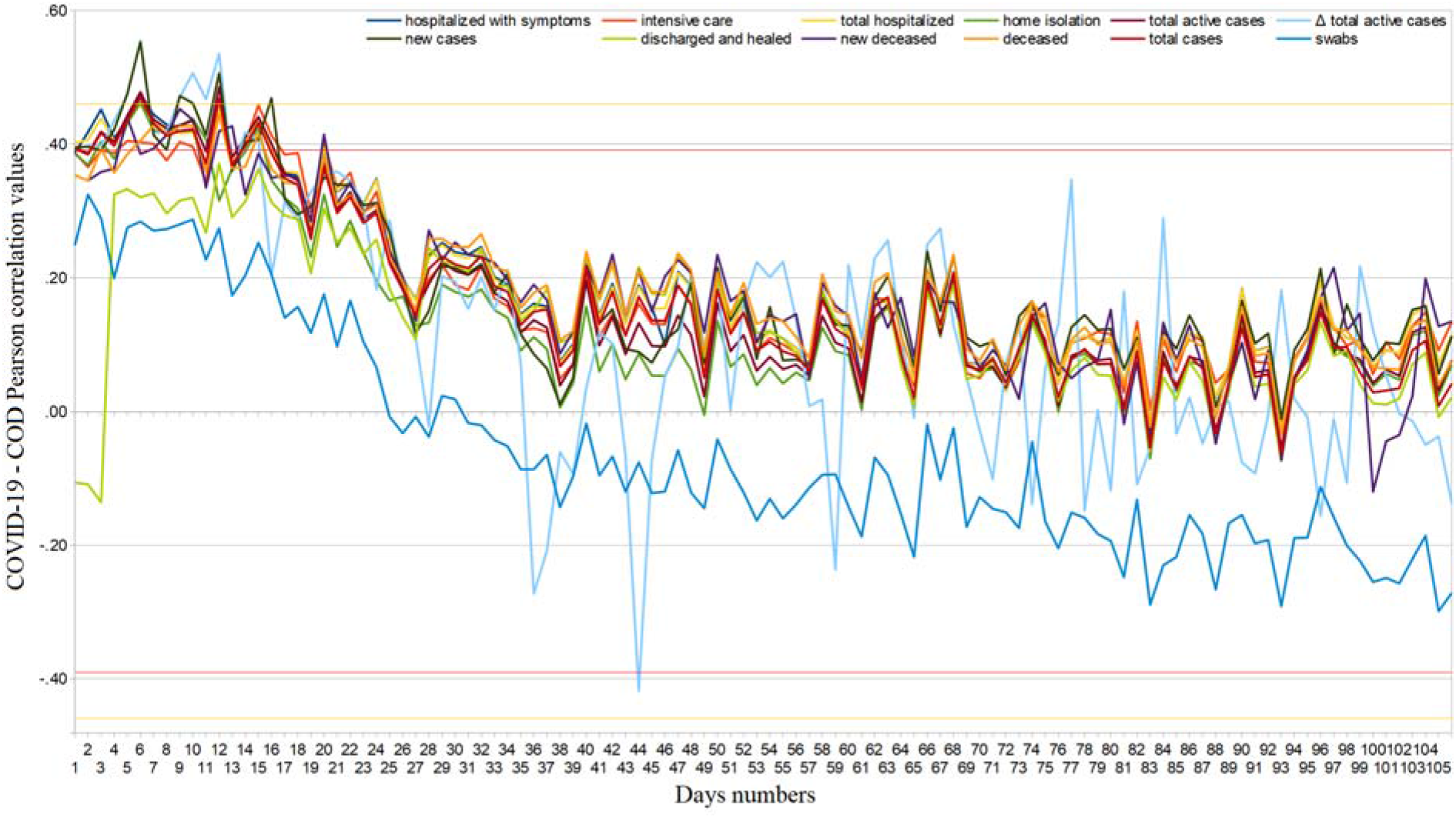
COVID-19 regional cumulative Web interest – official data Pearson’s correlation trends from February 24 to June 10, 2020.

### Hygiene and prevention Web interest

Web search interest related to disinfectants went from *ASV* = 5.14 (95% *CI:* [4.90, 5.38]) in the period from January 1 to February 19, 2020, to *ASV* = 11.09 (95% *CI:* [8.56, 13.63]) in the period from February 20 to June 10, 2020, i.e. it underwent an average increase of 115.8*%* (95% *CI:* [115.3, 116.3]). Considering the same two periods, Web interest related to medical masks and gloves went from *ASV* = 3.62 (95% *CI:* [2.88, 4.36]) to *ASV* = 36.25 (95% *CI:* [29.97, 42.52]), that is, it underwent an average increase of 901.2*%* (95% *CI:* [850.3, 952,2]). From February 20 to June 10, 2020, approximately 12.6% of hygiene Web interest was made up of searches related to COVID-19 (95% *CI:* [11.0, 14.2]), with an almost constant trend until around May 20 [Figure 4]. Considering daily Web searches, we found significant COD correlation values in very few isolated cases [Figure 5]. Finally, regarding cumulated Web searches, we highlighted significant positive COD correlations from February 26 to March 7, 2020, with all fields except “home isolation” and “swabs” (*ρ_best_* = .43, 95% *CI:* [. 42, .44],*p* = .07) [Figure 6]. Changes in the distribution of interest were more important than those on generic news (*ASV* = 86,*SD* = 9; *ASV* = 85,*SD* = 9) [Table 1].

**Figure 4.**
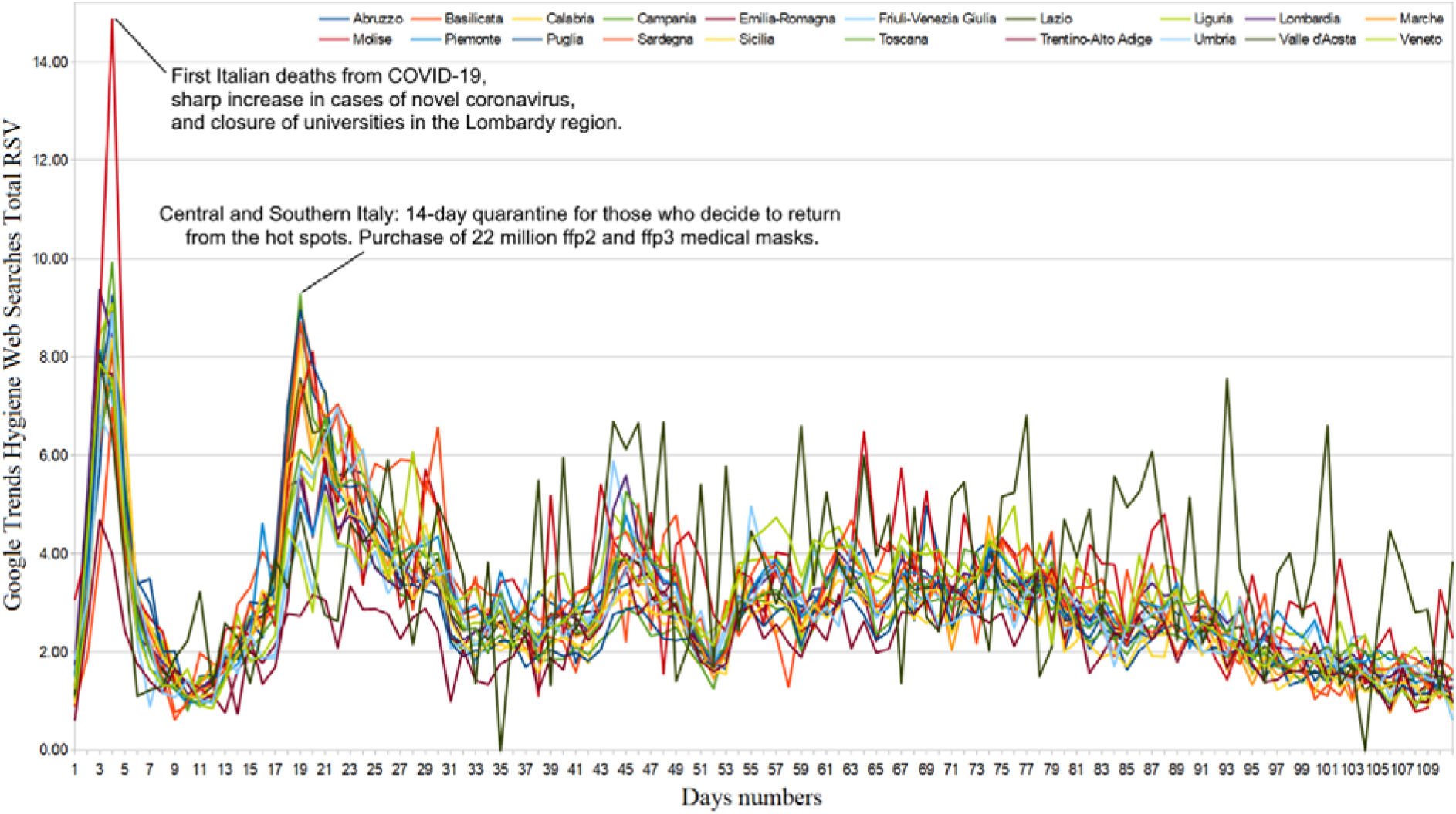
Hygiene and prevention regional Web interest from February 21 to June 10, 2020.

**Figure 5.**
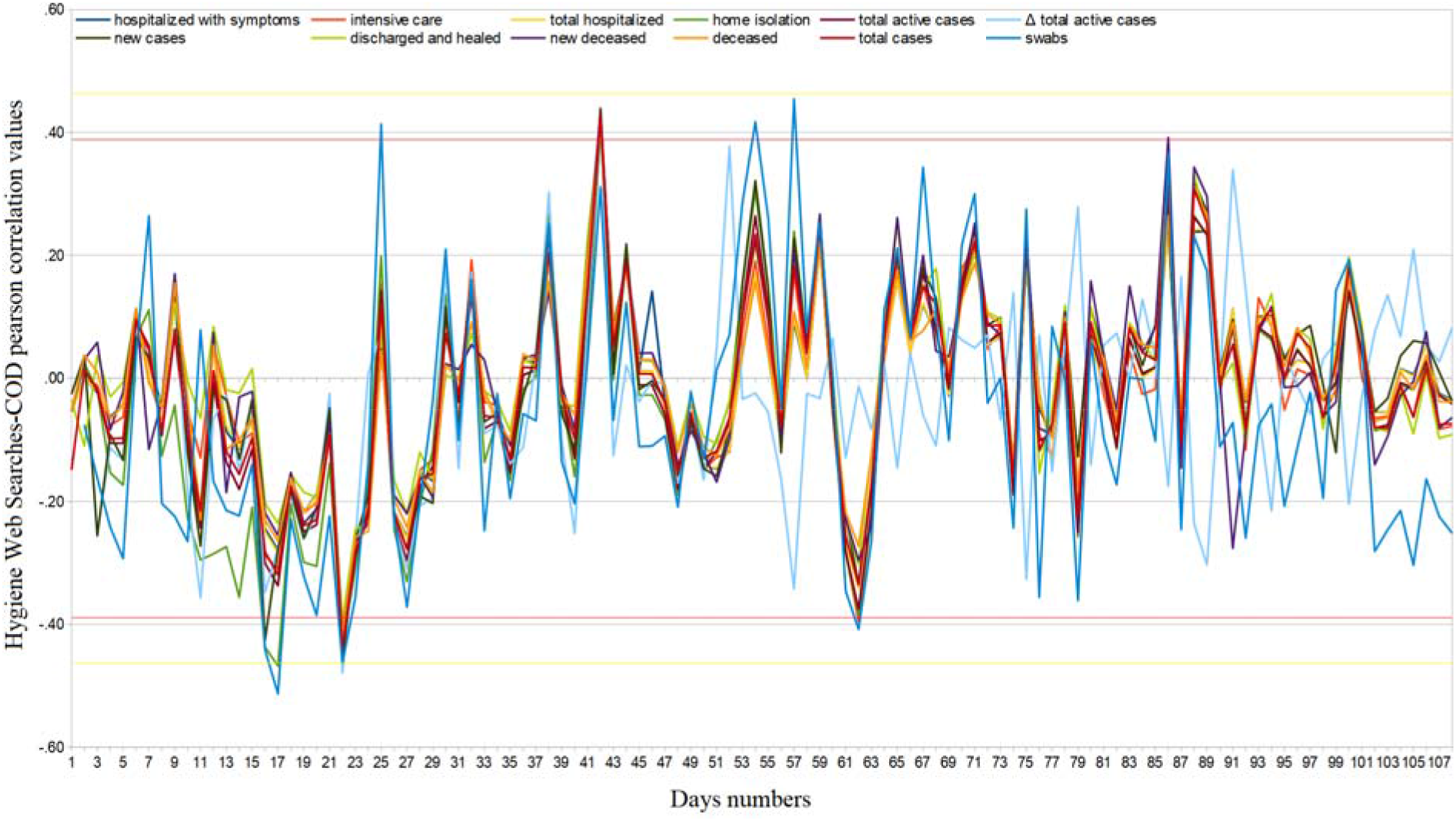
Hygiene daily regional Web interest – official data Pearson’s correlation trends from February 24 to June 10, 2020.

**Figure 6.**
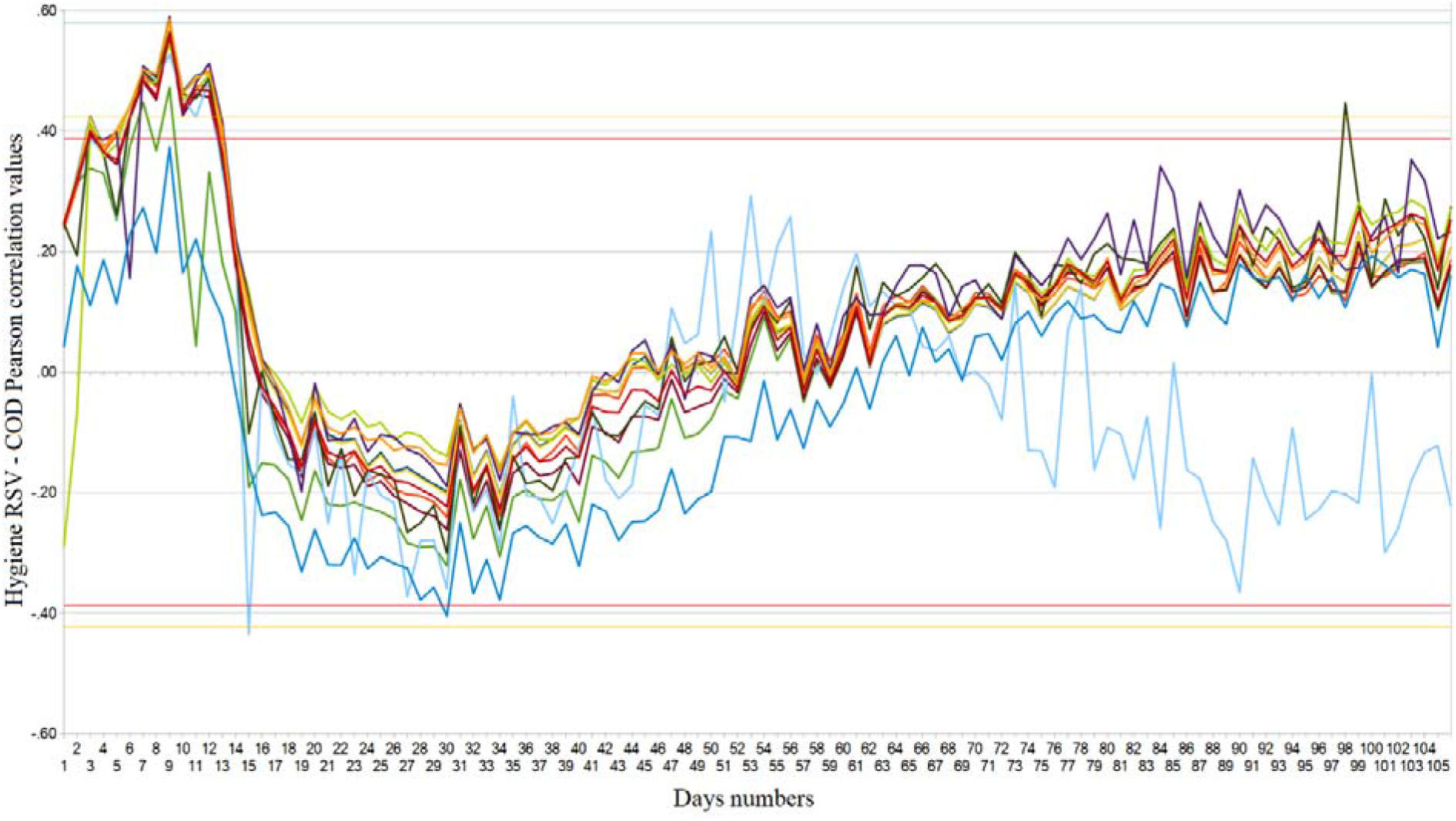
Hygiene regional cumulative Web interest – COVID-19 official data Pearson’s correlation trends from February 24 to June 10, 2020.

### Emotive-psychological well-being Web interest

Regarding the Web emotional response to COVID-19, we obtained the following results: nationally, comparing the average weekly RSVs of the two periods January 1, 2018– December 31, 2019 and January 1, 2020–June 10, 2020, we observed decreases of 3.1*%* and 16.1*%* for generic Web searches related to stress and depression, respectively (95% *CI:* [*−*3.0, *−*2.9],95% *CI:* [*−*16.2, *−*16.0]) and increases of 8.1*%* (95% *CI:* [8.0, 8.2]) and 20.5*%*(95% *CI:* [20.4, 20.6]) of those inherent in fear and anxiety (we point out that the trend related to anxiety had already been growing since December 2019) [Figure 7]. In the second period, the latter searches had a slightly increasing daily trend expressed by the equation *y = mx*, with *m* = 0.04, 95% *CI:* [0.00, 0.08]; this shows that general anxiety has not diminished over time, in contrast with the trends in numbers of new, active, and serious cases, and new deceased. From February 20 to June 10, 2020, 4.1% of the Web interest in negative emotions was made up of searches related to COVID-19 (95% *CI:* [3.6, 4.5]). In the same period, the Web interest in negative emotions explicitly related to novel coronavirus made up about 1.4*%* that related to COVID-19 (95% *CI:* [1.2, 1.5]). Regarding the latter, it was only possible to look for COD correlations with the cumulative Web interest because the data on the daily Web interest were too uncertain. We found significant cyclically positive COD correlations throughout the investigated period (Figure 8). To support this, Web interest in negative emotions has had extremely different distributions across regions (*ASV* = 55,*SD* = 41).

**Figure 7.**
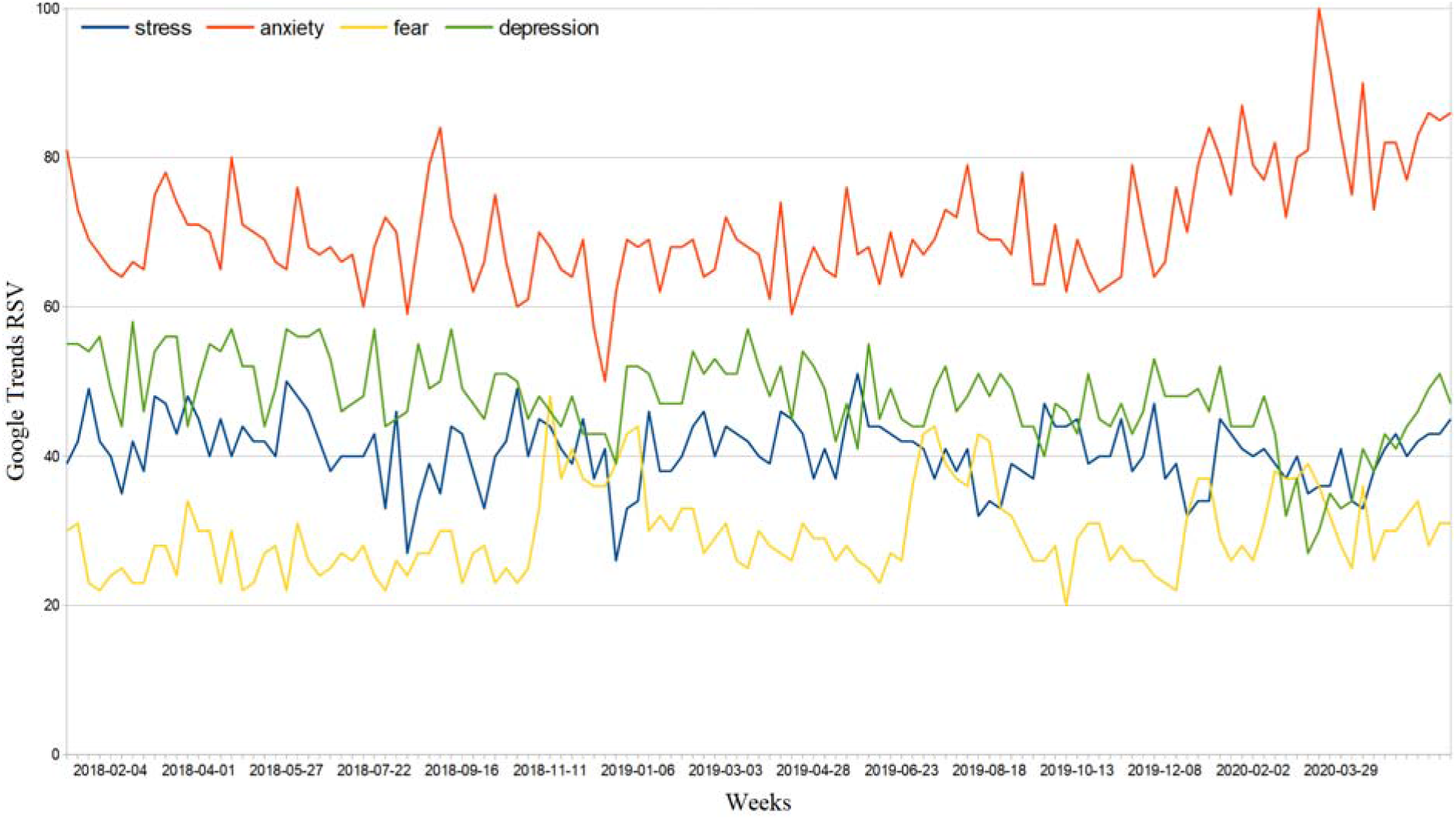
Italy Web interest in negative emotions from January 1, 2018, to June 10, 2020.

**Figure 8.**
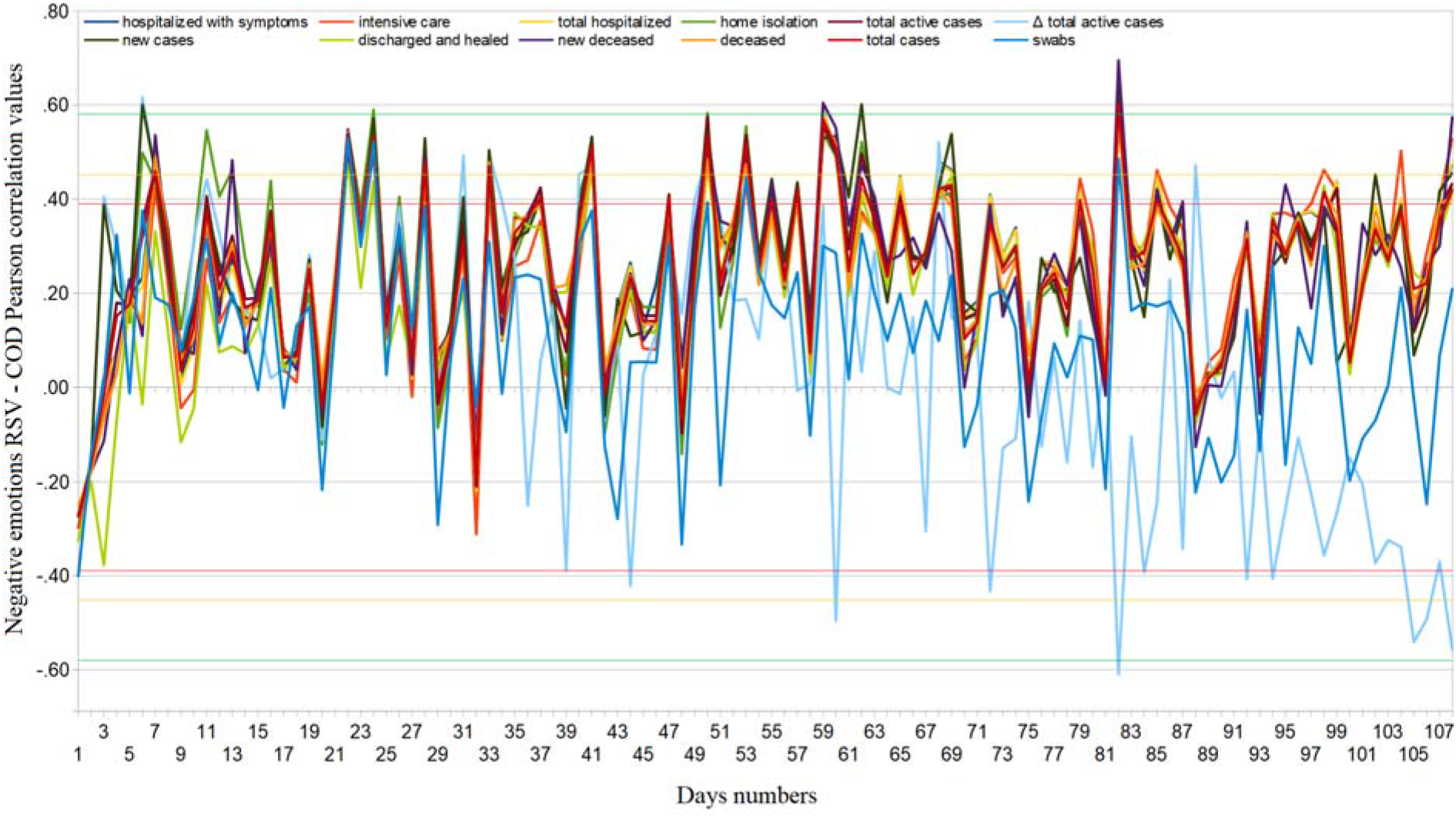
Regional cumulative Web interest in negative emotions related to novel coronavirus – COVID-19 official data Pearson’s correlation trends from February 24 to June 10, 2020.

## Discussion

The quantitative investigation of Web users’ interests in a crisis moment is a priority aspect to test the effectiveness of online information and the perception of risk by the population. This is the first study to carry out such an analysis in Italy, one of the countries worst affected by the COVID-19 pandemic. Our results show that interest in the novel coronavirus has exploded only in the presence of two contemporary phenomena: i) the presence of the virus in the nation, and ii) a significant number of cases. Even the state of emergency declared on January 31, 2020, due to a case of two Chinese tourists visiting Italy while infected with COVID-19, caused a very moderate Web interest for a limited duration [10]. We also highlighted a disparity between online searches and the actual perception of the danger: in fact, as shown in other studies and reported by national and international newspapers, the first episodes of racism against Chinese people and the so-called “coronavirus psychosis” [6, 11, 12]. Therefore, we believe it is plausible that the analysis of Web interest in Italy shows genuine interest only if the issue exceeds a certain severity threshold. We believe that the climate of uncertainty created by the press and by the many conflicting opinions of scientists negatively influenced the perception of the risk linked to COVID-19 in the Italian population. In particular, some have compared it to an influence, while others rightly denoted the dangers and critical issues also related to the lack of reliable information [13, 14]. When the problem became national, we recorded two peaks in the Web queries (February 27, 2020 and around March 14, 2020). Significant correlations between Web interest and regions with multiple cases rarely occurred in the initial phase, i.e. Web interest in the novel coronavirus has been linked only to its presence on national soil and nothing more. Since the aforementioned peaks had a maximum duration of about 7 days, we believe that online information is effective only in that time frame. The regional Web interest in this topic was very similarly distributed. This leads us to propose two considerations: i) the online information on a problem in its initial phase can heavily influence the thinking of the Italian population; ergo, it is important that those who present information online are clear right away, and ii) the online information on a problem in its initial phase can have a deep impact on the problem’s evolution.

Positive data emerged from Web interest in hygienic precautions, such as disinfectants and masks. In fact, the respective increases of approximately 115% and 901% compared to the periods preceding the virus signals a drastic change in netizens’ habits to face the pandemic. We must weigh such a result on the fact that the mean value of these queries corresponds to approximately 13% of all novel coronavirus-related queries. The lack of long-term correlations with the number of cases and the low variance of the data suggest that the interest was similarly distributed among all regions; this helped to avoid the spread of the epidemic nationwide [Table 1]. Even in this case, we have seen two peaks in queries and then a waning interest. This does not mean that the actual interest in hygiene has decreased, since disinfectants, masks, and gloves have experienced a substantial increase in sales [15].

General interest in negative feelings such as stress and depression fell during the lockdown (around 3% and 16%, respectively). The RSVs of anxiety and fear increased by approximately 21% and 8%, respectively. Therefore, it is plausible that there has been a shift in interest towards the latter. Direct associations between these symptoms of distress and the novel coronavirus made up about 1% of total COVID-19 queries. The data had an incredibly significant variance and high linear correlation values, i.e. the interest was much more pronounced in some regions, such as Emilia Romagna, Piedmont, Campania, Lombardy, and Puglia [Table 1]. The inconsistent trend of the correlation between cumulative Web searches and COVID-19 official data shown in Figure 8 suggests that the number of total searches was low, as it was easily influenced from day to day. The effects of the novel coronavirus and its lockdown on the mental health of the Italian population have been serious, as shown in other studies [7]. Therefore, even a small amount of these types of queries can mean a lot for mental disorders.

## Limitations

Web searches provide quantitative values only for users who use the Internet to obtain information on certain topics. All the queries investigated were collected from the Google search engine.

## Conclusions

Italian netizens showed a marked interest in the COVID-19 pandemic only when this became a direct national problem. In general, Web searches have rarely been correlated with the number of cases per region; we conclude that the danger, once it arrived in the country, was perceived similarly in all regions. We can state that the period of maximum effectiveness of online information, in relation to this type of situation, is limited to 3-4 days from a specific key event. If such a scenario were to occur again, we suggest that all government agencies focus their Web disclosure efforts over that time. Despite this, we found cyclical correlations with research related to negative feelings such as anxiety, depression, fear, and stress. Therefore, to identify mental and physical health problems among the population, it suffices to observe slight variations in the trend of related Web queries.

## Data Availability

All the data related to this study are presented in the manuscript.

## Acknowledgment

None

## Data availability

All the data related to this study are presented in the manuscript.

